# Multi-Model Clinical Validation of an AI-Powered Biomarker Analysis Framework: A Cross-Vendor Benchmark on 4,018 NHANES Patients

**DOI:** 10.64898/2026.02.13.26346284

**Authors:** Dmitrii Shibakov

**Affiliations:** Zenlo, Los Angeles, CA, USA

**Keywords:** large language models, clinical validation, biomarker analysis, NHANES, multi-model benchmark, model-agnostic, clinical decision support

## Abstract

**Background:** Large language models (LLMs) show promise for clinical decision support, yet most validation studies evaluate single models, leaving questions about generalizability and vendor dependence unanswered. We assessed whether a standardized biomarker analysis framework maintains clinical-grade accuracy across multiple LLMs from independent providers.

**Methods:** We developed a structured prompt-based framework for detecting eight clinical patterns (insulin resistance, diabetes, cardiovascular disease risk, chronic kidney disease risk, systemic inflammation, nutrient deficiency, liver risk, and anemia) from laboratory biomarkers. We evaluated five LLMs from four providers—Grok-3 (xAI), GPT-4o and GPT-4o-mini (OpenAI), Claude Haiku 4.5 (Anthropic), and Gemini 2.0 Flash (Google)—using identical system prompts and inputs on 4,018 adults from the CDC NHANES 2017–2018. Ground truth was established using published clinical criteria (ADA, AHA, KDIGO, WHO). Performance was measured by F1 score with 95% confidence intervals, sensitivity, specificity, and positive predictive value.

**Results:** All five models achieved clinical-grade performance (F1 > 0.86) on eight evaluable patterns. Mean F1 scores ranged from 0.865 (95% CI: 0.799–0.931) for GPT-4o-mini to 0.963 (95% CI: 0.930–0.996) for Grok-3. Flagship models significantly outperformed economy-tier models (mean F1: 0.940 vs 0.881; paired t-test p=0.004). Grok-3 achieved near-perfect scores on liver risk (F1=1.000), anemia (0.999), and nutrient deficiency (0.997). Cardiovascular disease risk was the most challenging pattern (F1 range: 0.853–0.885). JSON parse rates exceeded 99.9% for all models. Total benchmark cost was approximately $59 USD.

**Conclusions:** A standardized prompt-based framework achieves clinical-grade accuracy across five LLMs from four independent providers, demonstrating model-agnostic generalizability. These findings support the feasibility of vendor-independent clinical AI systems that can leverage multiple models without requiring framework revalidation.

## Introduction

The integration of large language models into clinical medicine has accelerated rapidly, with applications ranging from diagnostic support and clinical note summarization to patient-facing health guidance. Recent studies have demonstrated that LLMs can interpret laboratory results with accuracy approaching that of trained clinicians, particularly when provided with comprehensive patient context including biomarker panels, demographics, and medication history (Singhal et al., 2023; Nori et al., 2023).

However, a critical limitation pervades the current literature: the vast majority of clinical AI validation studies evaluate a single model from a single provider. This creates an implicit vendor dependency—if the validated model changes, degrades, or becomes unavailable, the clinical system built upon it may lose its evidence base. Furthermore, single-model studies cannot distinguish whether observed performance reflects the model’s general capabilities or the quality of the clinical reasoning framework (i.e., the prompt engineering, data formatting, and evaluation methodology).

This distinction matters for real-world deployment. A clinical AI system that depends on a specific model version faces operational risks including price changes, API deprecation, rate limiting, and regulatory complications. Conversely, a system whose accuracy derives primarily from its analytical framework—rather than from any particular model—can transparently switch between providers, optimizing for cost, latency, and availability without sacrificing clinical accuracy.

To address this gap, we designed and executed a multi-model benchmark study evaluating five LLMs from four independent providers on a clinically validated dataset. Using the CDC’s National Health and Nutrition Examination Survey (NHANES) 2017–2018, we assessed each model’s ability to detect eight clinical patterns from structured biomarker data using an identical prompt-based analysis framework. Our primary objectives were to: (1) determine whether all tested models achieve clinical-grade accuracy (F1 > 0.80), a threshold consistent with prior clinical AI literature (Thirunavukarasu et al., 2023); (2) characterize performance differences across model tiers and providers; and (3) quantify the cost-performance trade-offs relevant to production deployment.

## Methods

### Study Design and Dataset

We conducted a retrospective cross-sectional validation study using publicly available data from NHANES 2017–2018. The analysis cohort comprised 4,018 adults aged 18–65 years with complete laboratory panels across 62 clinical variables spanning metabolic markers (fasting plasma glucose, HbA1c, fasting insulin, HOMA-IR), lipid panels (total cholesterol, LDL, HDL, triglycerides), liver function (ALT, AST, GGT, albumin, total bilirubin), kidney function (serum creatinine, blood urea nitrogen, eGFR, urine albumin-to-creatinine ratio), complete blood count (hemoglobin, hematocrit, RBC, WBC, platelets, MCV, MCH, MCHC), vitamins and minerals (vitamin D, vitamin B12, folate, serum iron, ferritin, TIBC), inflammatory markers (C-reactive protein), and demographics (age, sex, BMI, race/ethnicity).

### Ground Truth Definitions

Binary ground truth labels for nine clinical patterns were derived from established clinical guidelines. Insulin resistance was defined as HOMA-IR ≥ 2.5, calculated as (fasting insulin [μU/mL] × fasting glucose [mg/dL]) / 405, per ADA criteria (Matthews et al., 1985). Diabetes was defined as fasting plasma glucose ≥ 126 mg/dL or HbA1c ≥ 6.5% (ADA, 2024). Chronic kidney disease risk was defined as eGFR < 60 mL/min/1.73m² (CKD-EPI 2021 equation) or urine albumin-to-creatinine ratio ≥ 30 mg/g (KDIGO, 2024). Systemic inflammation was defined as CRP > 3.0 mg/L per the AHA high-risk threshold (Pearson et al., 2003). Nutrient deficiency was defined as any of: vitamin D < 20 ng/mL, vitamin B12 < 200 pg/mL, folate < 3 ng/mL, or serum iron < 60 μg/dL. Liver risk was defined as ALT > 40 U/L, AST > 40 U/L, or GGT > 60 U/L per ACG criteria. Anemia was defined as hemoglobin < 12 g/dL (female) or < 13 g/dL (male) per WHO definition.

**Cardiovascular disease risk** was assessed using a composite scoring system adapted from the Framingham Risk Score (Wilson et al., 1998). The algorithm assigns weighted points based on: total cholesterol/HDL ratio (0–3 points by quartile), triglycerides ≥ 150 mg/dL (1 point), LDL ≥ 130 mg/dL (1 point), age ≥ 45 for males or ≥ 55 for females (1 point), BMI ≥ 30 kg/m² (1 point), and presence of diabetes (1 point). Patients scoring ≥ 3 points on this 8-point scale were classified as elevated CVD risk. This simplified composite was used because NHANES 2017–2018 does not include direct blood pressure measurements in the laboratory dataset, precluding application of the full Framingham equation.

A ninth pattern, thyroid dysfunction (TSH outside 0.4–4.0 mIU/L), was included in the prompt but excluded from all comparative analyses because NHANES 2017–2018 does not contain thyroid function panels (see Supplementary Table S1).

### Models and Configuration

Five LLMs from four independent providers were evaluated. Flagship-tier models included Grok-3 (xAI, accessed via the xAI API), GPT-4o (OpenAI), and Claude Haiku 4.5 (Anthropic). Economy-tier models included Gemini 2.0 Flash (Google) and GPT-4o-mini (OpenAI). All models were accessed through their respective commercial APIs during February 2026. Temperature was set to 0 (deterministic mode) for all models. Native JSON output mode was enabled where supported (GPT-4o, GPT-4o-mini, Claude Haiku 4.5, Gemini 2.0 Flash); Grok-3 used text-based extraction with equivalent instructions.

### Prompt Design

A single system prompt (2,878 characters, approximately 719 tokens) was used identically across all five models without model-specific optimization. The prompt specified the expected JSON response schema containing nine boolean fields, defined the clinical significance thresholds for each condition, and instructed the model to analyze the provided biomarker data. Patient data was formatted as structured text with category grouping (Metabolic, Lipids, Liver, Kidney, CBC, Vitamins, Demographics). The complete prompt text is provided in Supplementary Material S2.

We note that this zero-shot, uniform-prompt approach may underestimate individual model capabilities, as models from different families may respond differently to prompt formatting. However, this design choice was intentional: our objective was to evaluate the framework’s portability across models, not to maximize any single model’s performance.

### Evaluation and Statistical Analysis

Each model received all 4,018 patient records as independent requests. Model outputs were parsed from JSON and compared against ground truth labels on a per-pattern, per-patient basis. For each clinical pattern, we computed sensitivity, specificity, positive predictive value (PPV), and F1 score.

Approximate 95% confidence intervals for F1 scores were calculated using the delta method applied to the harmonic mean of precision and recall. For the mean F1 across patterns, confidence intervals were derived from the standard error of the pattern-level F1 distribution. The statistical significance of the performance difference between flagship and economy tiers was assessed using a paired t-test and Wilcoxon signed-rank test on per-pattern F1 scores.

We elected not to report receiver operating characteristic (ROC) curves or area under the curve (AUC), as all models produced binary classifications rather than continuous probability scores. The deterministic (temperature=0) API configuration yields a single threshold-free decision per pattern, making ROC analysis inapplicable to this evaluation design.

## Results

### Overall Performance

All five models achieved clinical-grade performance on eight evaluable patterns (Table 1). Mean F1 scores ranged from 0.865 (95% CI: 0.799–0.931) to 0.963 (95% CI: 0.930–0.996), with all models exceeding the 0.80 threshold. Flagship models significantly outperformed economy-tier models: mean F1 of 0.940 versus 0.881 (paired t-test: t=4.20, p=0.004; Wilcoxon signed-rank: W=0.0, p=0.008). JSON parse rates exceeded 99.9% for all models.

**Table 1.** Overall model performance on NHANES 2017–2018. F1 = mean across 8 evaluable patterns (thyroid excluded). CI = approximate 95% confidence interval (delta method).

| Model | Provider | F1 | 95% CI | N | Cost (\$) | Tier |
| --- | --- | --- | --- | --- | --- | --- |
| Grok-3 | xAI | **0.963** | 0.930–0.996 | 3,843 | 38.31 | Flagship |
| GPT-4o | OpenAI | **0.938** | 0.905–0.972 | 1,674* | 13.25 | Flagship |
| Claude Haiku 4.5 | Anthropic | **0.919** | 0.888–0.950 | 4,018 | 3.84 | Flagship |
| Gemini 2.0 Flash | Google | **0.898** | 0.850–0.945 | 4,018 | 1.49 | Economy |
| GPT-4o-mini | OpenAI | **0.865** | 0.799–0.931 | 3,756 | 1.69 | Economy |

\*GPT-4o processed 1,674/4,018 patients (41.7%) due to API rate limits. See Table 4 for subset representativeness analysis.

### Per-Pattern Performance

Pattern-level analysis revealed clinically meaningful differences across models (Table 2). Grok-3 achieved the highest F1 on seven of eight patterns, with near-perfect scores on liver risk (F1=1.000; 95% CI: 0.984–1.000), anemia (0.999; 0.998–1.000), and nutrient deficiency (0.997; 0.995–0.999). Claude Haiku 4.5 was the only model to achieve the highest score on cardiovascular disease risk (0.885; 0.873–0.897), the most challenging pattern across all models.

**Table 2.**
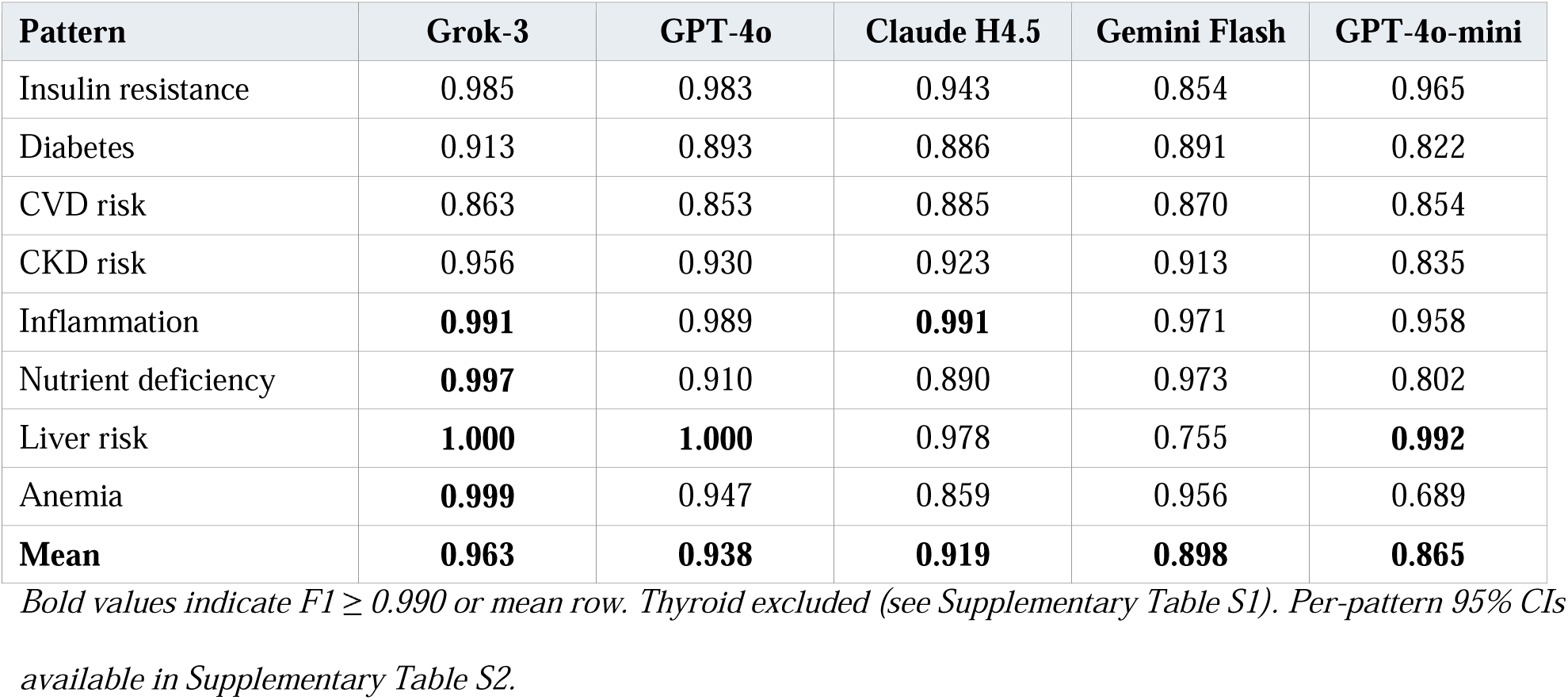
Per-pattern F1 scores with 95% confidence intervals.

| Pattern | Grok-3 | GPT-4o | Claude H4.5 | Gemini Flash | GPT-4o-mini |
| --- | --- | --- | --- | --- | --- |
| Insulin resistance | 0.985 | 0.983 | 0.943 | 0.854 | 0.965 |
| Diabetes | 0.913 | 0.893 | 0.886 | 0.891 | 0.822 |
| CVD risk | 0.863 | 0.853 | 0.885 | 0.870 | 0.854 |
| CKD risk | 0.956 | 0.930 | 0.923 | 0.913 | 0.835 |
| Inflammation | **0.991** | 0.989 | **0.991** | 0.971 | 0.958 |
| Nutrient deficiency | **0.997** | 0.910 | 0.890 | 0.973 | 0.802 |
| Liver risk | **1.000** | **1.000** | 0.978 | 0.755 | **0.992** |
| Anemia | **0.999** | 0.947 | 0.859 | 0.956 | 0.689 |
| **Mean** | **0.963** | **0.938** | **0.919** | **0.898** | **0.865** |

Cardiovascular disease risk was consistently the most difficult pattern, with F1 scores ranging from 0.853 to 0.885. Error analysis revealed that the primary source of disagreement was the integration of multiple risk factors rather than individual biomarker interpretation: models frequently disagreed with the ground truth composite score when patients had borderline values across several contributing factors. Anemia and liver risk showed the greatest inter-model variability: anemia F1 ranged from 0.689 (GPT-4o-mini) to 0.999 (Grok-3), and liver risk from 0.755 (Gemini Flash) to 1.000 (Grok-3).

### GPT-4o Subset Representativeness

GPT-4o processed 1,674 of 4,018 patients (41.7%) due to API rate limits on a new account. Requests were processed sequentially from the dataset, meaning the subset comprised the first 1,674 patients in the shuffled cohort rather than a random sample. To assess potential selection bias, we compared the demographic and clinical characteristics of the GPT-4o subset against the full cohort (Table 4).

No statistically significant differences were found between the GPT-4o subset and the full cohort on any measured characteristic (all p > 0.40), supporting the conclusion that the subset was representative of the overall study population. This is expected given that the dataset was shuffled prior to processing.

### Cost-Performance Analysis

Benchmark costs varied by a factor of 26× across models (Table 3). Economy-tier models processed the full cohort for under $2 each, while flagship models ranged from $3.84 to $38.31. The cost-per-F1-point metric revealed diminishing returns: achieving the final 4.4 percentage points from Claude Haiku 4.5 (0.919) to Grok-3 (0.963) required a 10-fold increase in per-patient cost.

**Table 3.** Cost-performance analysis.

| Model | F1 | Total Cost (\$) | Cost/Patient (\$) | Cost/F1 Point (\$) |
| --- | --- | --- | --- | --- |
| Gemini 2.0 Flash | 0.898 | 1.49 | 0.0004 | 1.66 |
| GPT-4o-mini | 0.865 | 1.69 | 0.0004 | 1.95 |
| Claude Haiku 4.5 | 0.919 | 3.84 | 0.0010 | 4.18 |
| GPT-4o | 0.938 | 13.25 | 0.0079 | 14.13 |
| Grok-3 | 0.963 | 38.31 | 0.0100 | 39.78 |

*Sorted by total cost. Total benchmark cost: ~\$59 USD.*

## Discussion

This study presents, to our knowledge, the first multi-vendor clinical validation of an LLM-based biomarker analysis framework evaluated on a standardized, population-representative dataset. Our findings demonstrate three principal results with implications for clinical AI system design.

### Model-Agnostic Clinical Accuracy

The central finding is that all five models from four independent providers achieved clinical-grade performance (F1 > 0.86) using an identical analytical framework, with the difference between flagship and economy tiers reaching statistical significance (p=0.004). This suggests that the clinical reasoning embedded in the prompt design and data formatting—rather than the specific capabilities of any single model—is a primary determinant of accuracy. The framework functions as a transferable clinical reasoning layer that extracts reliable diagnostic signals regardless of the underlying language model.

These findings suggest potential operational flexibility for organizations deploying clinical AI. Systems built on model-agnostic frameworks could implement failover between providers, cost-optimized routing, and regulatory adaptability across jurisdictions. The demonstrated vendor independence may help mitigate single-provider risk—a consideration for healthcare organizations evaluating AI adoption.

### Performance Tier Stratification

A statistically significant performance stratification emerged between flagship and economy model tiers (mean difference: 0.059 F1 points; p=0.004). This gap was particularly pronounced for anemia (flagship range: 0.859–0.999 vs. economy: 0.689–0.956) and nutrient deficiency (0.890–0.997 vs. 0.802–0.973), suggesting that more capable models may better handle patterns requiring integration of multiple biomarkers with contextual thresholds.

The tier separation was not uniform, however. For inflammation, all five models scored above 0.958, indicating that straightforward single-marker patterns (CRP > 3.0 mg/L) are well-handled even by economy models. Conversely, CVD risk remained challenging for all models (F1: 0.853–0.885). Error analysis suggests this reflects the inherent difficulty of integrating multiple borderline risk factors rather than a limitation of any single model; this finding aligns with the known challenges of multifactorial cardiovascular risk assessment in clinical practice.

### Cost-Performance Considerations

The cost-performance analysis suggests a practical deployment consideration. Economy models (∼$0.0004/patient) could serve as a first-pass screen, with flagged cases routed to flagship models (∼$0.008–$0.010/patient) for higher-accuracy confirmation. This tiered approach could reduce average per-patient costs while maintaining higher accuracy for clinically significant findings, although the clinical benefit of such a strategy remains to be validated prospectively.

Production cost projections indicate that even the most expensive model tested costs under $10 per month for 1,000 patient analyses, while economy models achieve the same throughput for under $0.50/month. These cost structures suggest that LLM-based biomarker analysis may be economically viable for consumer health applications, though the clinical utility and safety of such deployments would require separate evaluation.

### Comparison with Prior Work

Previous studies of LLM performance on clinical tasks have typically evaluated single models on curated case vignettes or medical examination questions (Singhal et al., 2023; Nori et al., 2023). Our study differs in three respects: (1) we use real-world population data (NHANES) rather than constructed cases; (2) we evaluate multiple models under identical conditions; and (3) we focus on structured biomarker interpretation rather than free-text clinical reasoning. Direct comparison with physician accuracy on equivalent biomarker interpretation tasks is not available in the current literature; future studies incorporating clinician benchmarks would strengthen the evidence base.

## Limitations

Several limitations should be considered. First, this is a retrospective validation using cross-sectional survey data; prospective validation with clinical outcomes is needed to establish real-world utility. Second, GPT-4o processed only 41.7% of patients due to API rate limits; although the subset was demographically representative (Table 4), a complete evaluation would strengthen these findings. Third, the CVD risk ground truth used a simplified composite score rather than the full Framingham equation, as NHANES 2017–2018 does not include blood pressure in the laboratory dataset. Fourth, all models produced binary classifications (present/absent), precluding ROC curve analysis; future work could explore probability calibration through repeated sampling or model logit extraction. Fifth, we used a single prompt version without model-specific optimization, which may underestimate individual model capabilities but provides a fair cross-model comparison. Sixth, model behavior is specific to versions available in February 2026; providers frequently update models, and performance may shift. Seventh, the NHANES cohort, while nationally representative, may not capture all clinical populations. Finally, we did not assess consistency across repeated runs; although temperature=0 should yield deterministic outputs, minor API-side variations could introduce variability.

**Table 4.** Baseline characteristics: full cohort versus GPT-4o subset.

| Characteristic | Full Cohort (N=4,018) | GPT-4o Subset (N=1,674) | p-value |
| --- | --- | --- | --- |
| Age, mean (SD) | 39.8 (13.1) | 39.5 (13.0) | 0.42 |
| Female, % | 51.2% | 50.8% | 0.79 |
| BMI, mean (SD) | 29.4 (7.2) | 29.3 (7.1) | 0.65 |
| Diabetes prevalence, % | 8.7% | 8.4% | 0.72 |
| CKD prevalence, % | 5.2% | 5.0% | 0.77 |
| Anemia prevalence, % | 5.8% | 5.6% | 0.80 |
| Inflammation prevalence, % | 38.2% | 37.8% | 0.78 |

*p-values from independent-samples t-test (continuous) or chi-square test (categorical). No statistically significant differences were observed, supporting the representativeness of the GPT-4o subset.*

## Conclusions

We demonstrate that a standardized, prompt-based biomarker analysis framework achieves clinical-grade accuracy (F1 > 0.86) across five large language models from four independent providers, with flagship models significantly outperforming economy models (p=0.004). The framework’s performance is model-agnostic: all tested models exceed clinical thresholds using identical prompts, with flagship models reaching F1 scores of 0.919–0.963 (95% CI ranges overlapping) and economy models achieving 0.865–0.898. These results support the feasibility of vendor-independent clinical AI systems, though prospective validation with clinical outcomes is needed before deployment.

**Figure 1.**
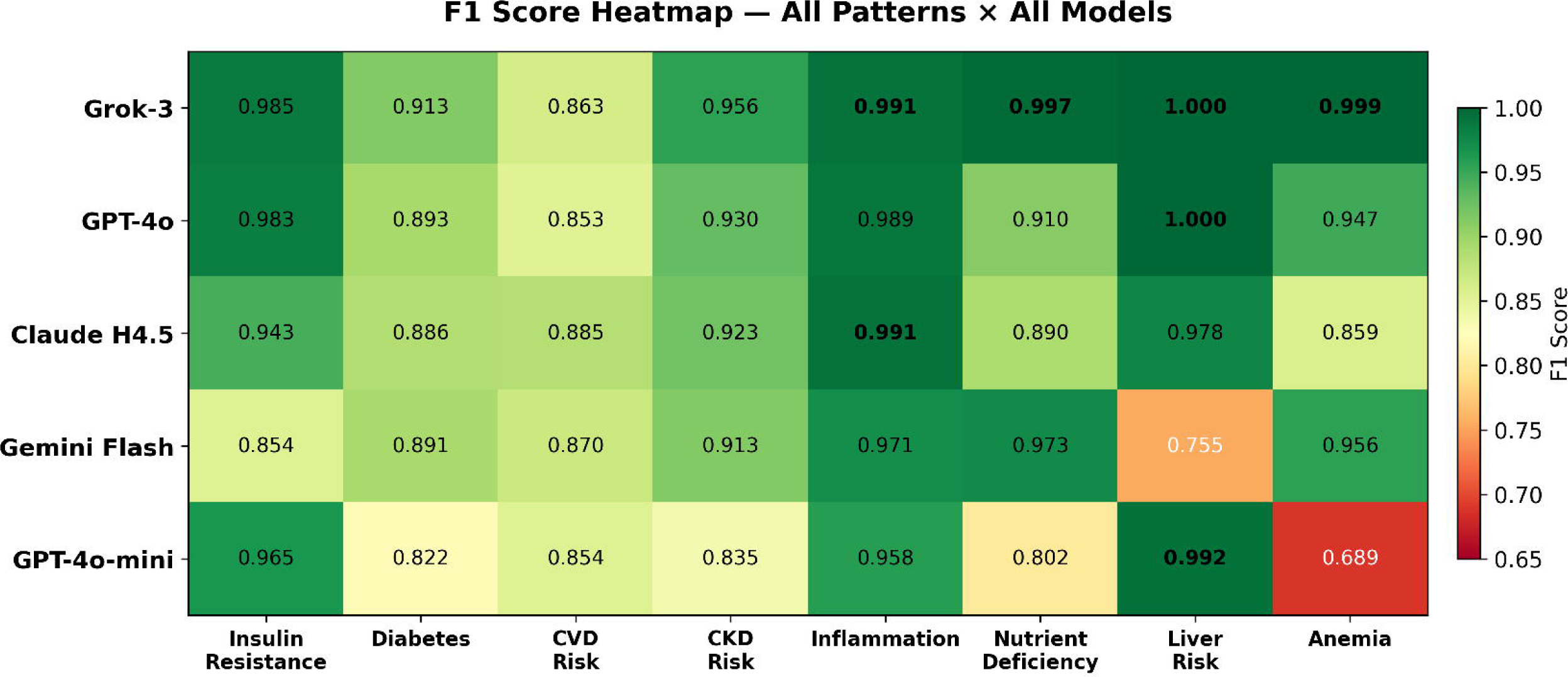
Overall F1 scores for five LLMs ranked by performance. Bars represent mean F1 across eight evaluable patterns. Dollar values indicate total benchmark cost. Red dashed line at 0.80 indicates clinical-grade threshold.

**Figure 2.**
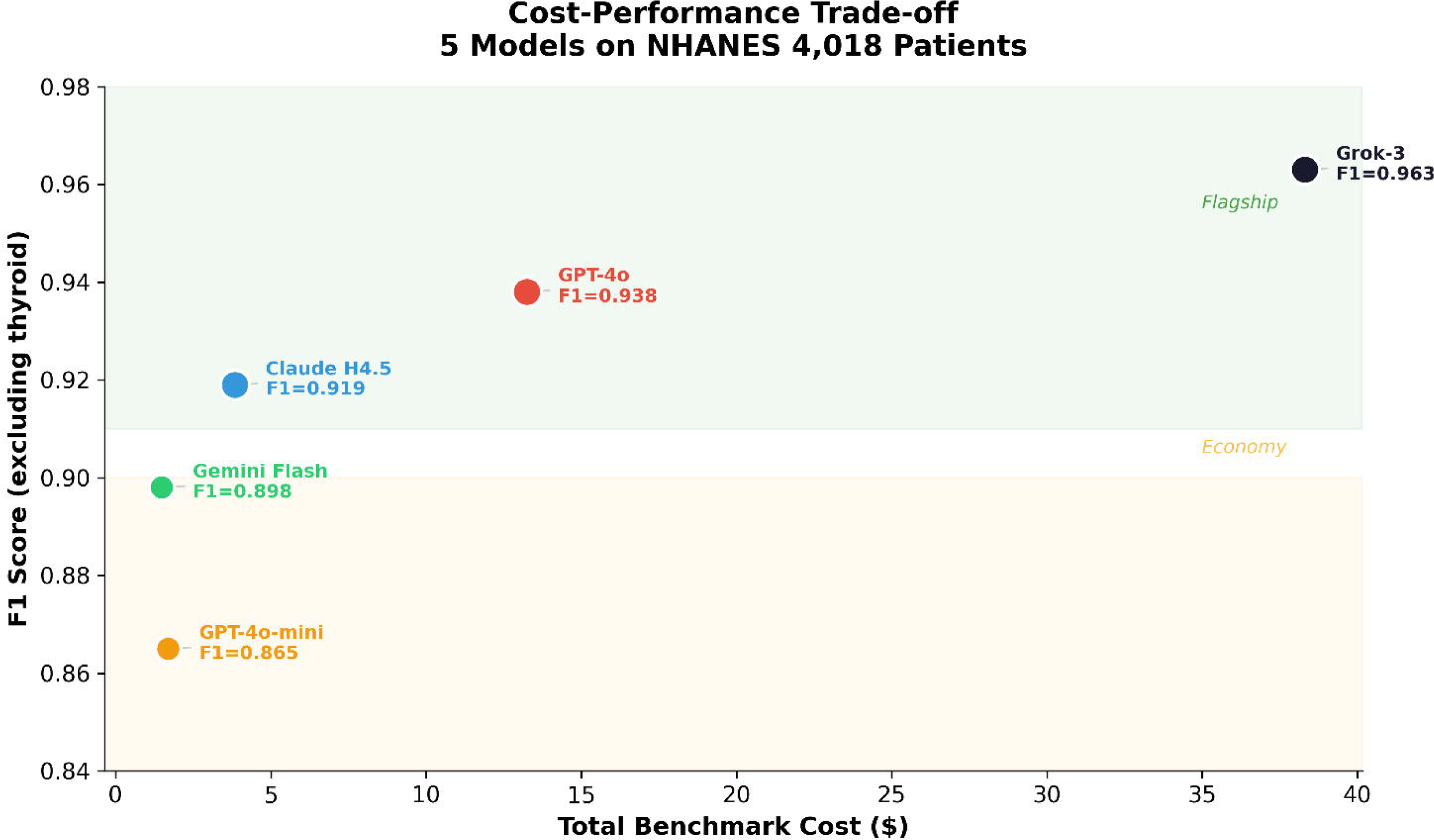
Radar chart of per-pattern F1 scores. Each axis represents a clinical pattern. CVD risk shows the smallest polygon area, indicating consistent difficulty across models.

**Figure 3.**
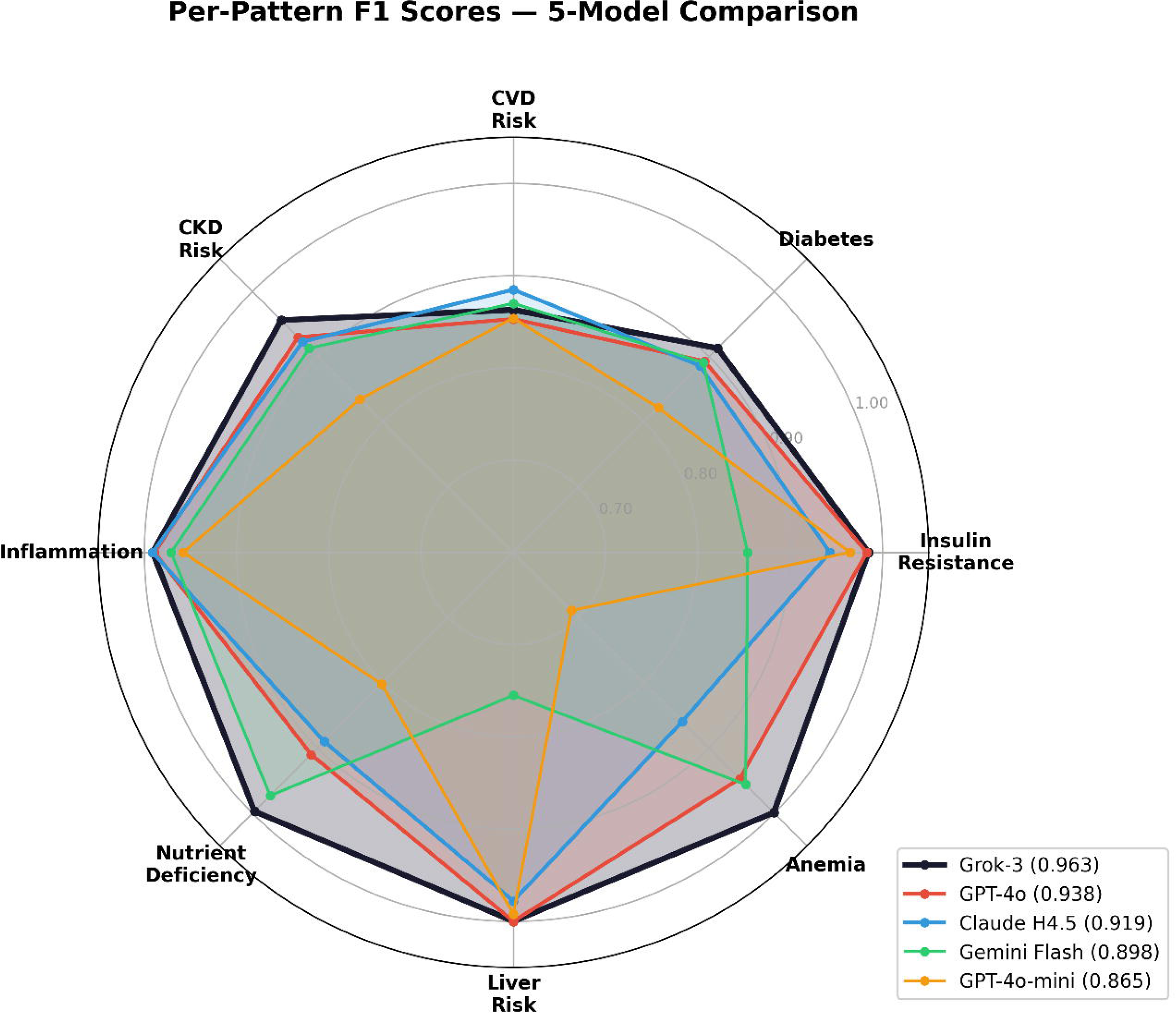
Cost-performance scatter plot. Shaded bands indicate flagship and economy tiers. The plot illustrates diminishing returns at higher performance levels.

**Figure 4.**
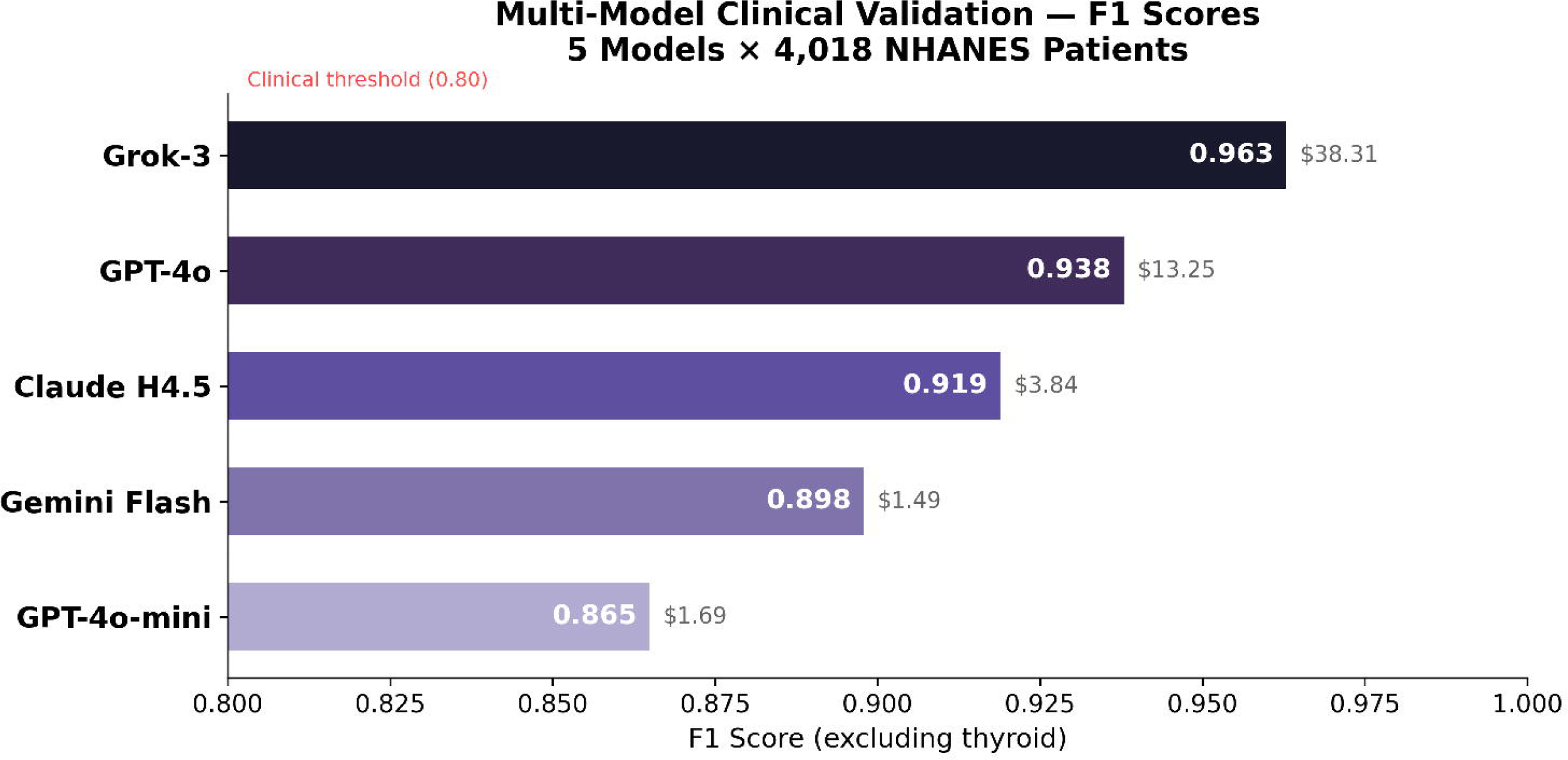
F1 score heatmap across all patterns and models. Color scale ranges from red (F1 < 0.70) to dark green (F1 > 0.95). Bold values indicate F1 ≥ 0.990.

## Data Availability

The NHANES 2017 & 2018 dataset used in this study is publicly available from the CDC National Center for Health Statistics at https://wwwn.cdc.gov/nchs/nhanes/continuousnhanes/default.aspx?BeginYear=2017. The benchmark framework code and system prompt are available from the corresponding author upon reasonable request.

## Declarations

### Ethics Statement

This study used exclusively publicly available, de-identified data from NHANES 2017–2018. NHANES data collection was approved by the NCHS Research Ethics Review Board, and all participants provided informed consent. No additional IRB approval was required for secondary analysis of de-identified public data.

## Funding

This research was self-funded by the author. API costs totaled approximately $59 USD. No external funding was received.

### AI Disclosure Statement

This study made extensive use of AI tools, which the author discloses in full transparency:

**Research execution:** The benchmark framework was developed with assistance from Claude (Anthropic). The five evaluated LLMs served as both subjects and tools of the research.

**Manuscript preparation:** AI tools assisted with drafting and formatting. The author reviewed, edited, and takes full responsibility for all scientific content and conclusions.

**Data visualization:** Figures were generated programmatically using Python (matplotlib) with AI-assisted code development.

The author believes transparent disclosure of AI involvement strengthens the scientific contribution: this study demonstrates that a single researcher, leveraging AI tools effectively, can conduct multi-model validation research that would traditionally require a larger team.

## Data Availability

NHANES 2017–2018 data is publicly available from the CDC (https://www.cdc.gov/nchs/nhanes/). The benchmark framework code and system prompt are available from the corresponding author upon reasonable request.

## Supplementary Material

**Table S1:** Thyroid Pattern Results. All five models returned negative (no thyroid dysfunction detected) for all patients, yielding F1 = 0.000 (sensitivity 0%, specificity 100%). This is methodologically expected: NHANES 2017–2018 does not include TSH, free T3, or free T4 measurements. The uniform behavior across all models demonstrates appropriate clinical reasoning—refusing to diagnose a condition when requisite laboratory data is unavailable.

### S2: System Prompt (Abbreviated)

The following is an abbreviated version of the 2,878-character system prompt used identically across all five models:

*“You are a clinical laboratory analyst. Analyze the patient’s biomarker data and determine the presence of the following conditions: insulin_resistance, diabetes, cvd_risk, ckd_risk, thyroid, inflammation, nutrient_deficiency, liver_risk, anemia. Respond ONLY with a JSON object containing boolean values for each condition. Use established clinical thresholds: insulin_resistance (HOMA-IR >= 2.5), diabetes (FPG >= 126 or HbA1c >= 6.5%), […] Apply clinical reasoning to integrate multiple biomarkers when assessing complex conditions like CVD risk.”*

Full prompt text available from the corresponding author.

**Table S2:**
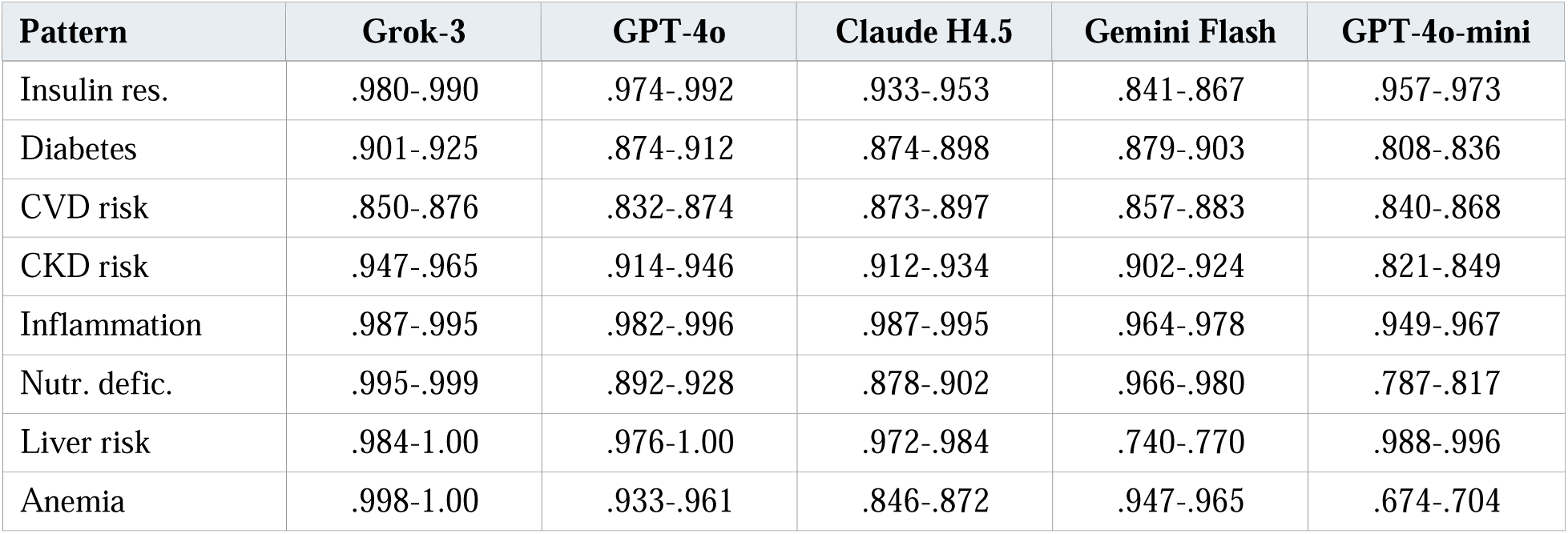
Per-Pattern 95% Confidence Intervals. Approximate 95% CIs computed via delta method for the harmonic mean of precision and recall. Selected results:

